# Predictive Biomarkers for the Responsiveness of Recurrent Glioblastomas to Activated Killer Cell Immunotherapy

**DOI:** 10.1101/2022.09.21.22280225

**Authors:** Sohyun Hwang, Jaejoon Lim, Haeyoun Kang, Ju-Yeon Jeong, Je-Gun Joung, Jinhyung Heo, Daun Jung, Kyunggi Cho, Hee Jung An

## Abstract

**Background:** Recurrent glioblastoma multiforme (GBM) is a highly aggressive primary malignant brain tumor that is resistant to existing treatments. Recently, we reported that activated autologous natural killer (NK) cell therapeutics induced a marked increase in survival of some patients with recurrent GBM.

**Methods:** To identify biomarkers that predict responsiveness to NK therapeutics, we examined immune profiles in tumor tissues using NanoString nCounter analysis and compared the profiles between 5 responders and 7 non-responders. Through a three-step data analysis, we identified three candidate biomarkers (*TNFRSF18, TNFSF4*, and *IL12RB2*) and performed validation with qRT-PCR. We also performed immunohistochemistry and an NK cell migration assay to assess the function of these genes.

**Results:** Responders had higher expression of many immune-signaling genes compared with non-responders, which suggests an immune-active tumor microenvironment in responders. The random forest model that identified *TNFRSF18, TNFSF4*, and *IL12RB2* showed a 100% accuracy (95% CI: 73.5%–100%) for predicting the response to NK therapeutics. The expression levels of these three genes by qRT-PCR were highly correlated with the NanoString levels, with high Pearson’s correlation coefficients (0.419 (*TNFRSF18*), 0.700 (*TNFSF4*), and 0.502 (*IL12RB2*)); their prediction performance also showed 100% accuracy (95% CI: 73.54%–100%) by logistic regression modeling. We also demonstrated that these genes were related to cytotoxic T cell infiltration and NK cell migration in the tumor microenvironment.

**Conclusions:** We identified *TNFRSF18, TNFSF4*, and *IL12RB2* as biomarkers that predict response to NK cell therapeutics in recurrent GBM, which might provide a new treatment strategy for this highly aggressive tumor.

## Background

Glioblastoma multiforme (GBM) is a highly malignant brain tumor that generally has a median survival of less than 20 months, while the 5-year survival rate is only 4%–5% [1]. Despite available aggressive standard therapies, such as radiation and temozolomide (TMZ), after maximal surgical resection, most patients experience recurrence, for which there are currently no effective treatments, and thus, these patients exhibit poor overall survival.

Although a remarkable survival benefit has been achieved in many solid tumors including melanoma and lung cancer from recent advances in cancer immunotherapy, clinical trials showed that immune checkpoint inhibitors, such as CTLA-4, PD-1, and PD-L1, failed to prolong survival in patients with glioblastoma [2-4]. The mechanism of resistance to these immunotherapeutic drugs is considered to involve both endogenous and exogenous factors. The endogenous factors are related to the GBM tumor cells [5] and include low tumor mutation burden (TMB) [6], extensive intra-tumoral heterogeneity [7], and a tumor microenvironment (TME) that inhibits the recruitment and function of T cells, such as through activation of the mitogen-activated protein kinase pathway, VEGF, and interleukin production. The exogenous factors are those that affect T cell activation. The most important external factor is the immune microenvironment. Infiltration of regulatory T (Treg) cells [8], myeloid-derived suppressor cells (MDSCs) [9], and tumor-associated macrophages (TAMs), which inhibit the antitumor activity of T cells and natural killer (NK) cells, might induce resistance to immunotherapy. Therefore, biomarkers that predict treatment sensitivity and that can be used to screen the population to determine who is suitable for immunotherapy are urgently needed to improve immunotherapy efficacy.

Recently, we performed a clinical trial that included 14 patients with recurrent GBM treated with autologous activated NK cells (AKCs) [10] and found that the progression-free and overall survival times were significantly increased in some patients in the treatment group, whereas the average survival of patients with recurrent GBM generally ranges from 5 to 10 months [11-14]. Of the 14 patients in the treatment group, 5 responded to the treatment, and survival was longer than 24 months.

In this study, to identify the characteristics of the tumor immune microenvironment profile in responders to AKC therapy and the factors that can predict treatment response, we examined the immune profiles of each patient’s tissue using NanoString nCounter analysis. We also performed immunohistochemistry to detect immune cells and analyzed the possible predictive biomarkers of response to this immune-cell therapy. In addition, we performed a functional study for the possible predictive biomarkers we identified.

## Materials and methods

### Patient samples

A previous clinical study was conducted with 14 patients with recurrent GBM as a single-arm, open-label, and investigator-initiated trial [10]. Autologous AKCs, which were expanded *ex-vivo* for 14 days and activated from peripheral blood mononuclear cells (PBMCs), were administered via intravenous injection 24 times at two-week intervals. Among the 14 patients, 5 showed a durable response and survived for over 2 years; the other nine patients survived less than 2 years after surgery, and MRI revealed progression during AKC treatment; these patients were classified as non-responders. A further study was performed on paraffin-embedded tissue samples of 12 patients (5 responders and 7 non-responders), which were obtained before AKC treatment and were of sufficient quantity and quality to be used in this study. Two board-certified pathologists (H.K and J.H.) reviewed the hematoxylin and eosin (H&E)–stained slides of all patients to evaluate tissue quality and to identify the tumor area to be analyzed. This study was approved by the Institutional Review Board of Bundang CHA Medical Center (#2012-12-172, #2021-01-024). Informed consent was obtained from each patient before the clinical trial.

### RNA extraction and NanoString nCounter analysis

The method for RNA extraction and NanoString nCounter analysis is described in the Supplementary Methods.

### Identification of genes significantly associated with treatment response and random forest modeling

To identify genes that are significantly associated with treatment response, we performed a three-step data analysis: area under the ROC curve (AUC), survival analysis, and random forest modeling. In the AUC analysis, we selected genes for which the AUC score was equal to or higher than 0.85. In the survival analysis, we selected genes for which the overall survival (OS) *p*-value or progression-free survival (PFS) *p*-value was less than 0.05. Lastly, we constructed random forest models with the identified significant genes by randomForest and care R packages. We used all default values for random forest parameters. The random forest model provided the importance scores, such as mean disease Gini score and mean disease accuracy, which represent gene importance for predictability. The mean decrease in Gini/accuracy scores was used to calculate how the model accuracy would decrease when each gene was excluded in model construction. A high score indicates greater importance for prediction. For performance measurement, the out-of-bag error rate of the random forest model, sensitivity, and specificity were calculated.

### Logistic regression modeling

To assess the prediction performance of selected genes, we applied a logistic regression model using the glm (generalized linear model) function in R along with the expression levels of NanoString nCounter. The prediction accuracy was evaluated based on leave-one-out cross-validation. In addition, we constructed the prediction model using logistic regression to distinguish responders from non-responders by RT-qPCR.

### RT-qPCR with TaqMan gene expression assays

RNAs (500 ng) were reverse transcribed using SuperScript™ IV VILO™ Master mix (Invitrogen, Waltham, MA, USA). RT-qPCR quantitation was performed using TaqMan™ Gene expression assays [FAM] (Applied Biosystems, Carlsbad, CA, USA) for *TNFSF4* (Hs01911853_s1), *TNFRSF18* (Hs00188346_m1), and *IL12RB2* (Hs00155486_m1) and TaqMan™ Gene expression Master Mix (Applied Biosystems, Foster City, CA, USA) on a Bio-Rad CFX96 Real-Time PCR Detection System (Bio-Rad). TBP (Hs00427620_m1) was used as a control to allow quantitation of relative gene expression in RNA samples. All PCR reactions were amplified in duplicate, and relative gene expression levels were calculated using the -ΔCt method.

### Immunohistochemistry and digital image analysis

Immunohistochemistry for CD3 (2GV6) (Roche Diagnostics International AG, Rotkreuz, Switzerland), CD8 (SP57) (Roche Diagnostics International AG, Rotkreuz, Switzerland), and CD68 (KP-1) (Roche Diagnostics International AG, Rotkreuz, Switzerland) was performed on formalin-fixed paraffin-embedded (FFPE) tissues from all 12 cases on a fully automated stainer (VENTANA BechMark ULTRA, Roche Diagnostics International AG, Rotkreuz, Switzerland). The secondary antibodies with horse radish peroxidase (HRP)-3,3′-diaminobenzidine tetrahydrochloride (DAB) were applied as instructed by the manufacturer (See Supplementary Table 1 for more details). For each immunohistochemical stain, the number of positive cells per mm^2^ was detected and calculated on QuPath [15]. Whole slide images were scanned at 20x using a digital slide scanner (APERIO AT2, Leica Biosystems, Nussloch, Germany). Tumor and non-tumor areas of the whole slide image of representative sections for each patient were annotated with QuPath.

### Transwell migration assay for NK cell migration to assess the function of the three genes

To assess whether these three genes were expressed in T cells and whether they were related to NK cell migration to the tumor, we performed a Transwell migration assay mimicking the tumor microenvironment. We detected the number of NK cells in the upper chamber that migrated to the lower chamber, which contained GBM cancer cells and T cells that either expressed or did not express these three genes. The human glioblastoma cell line U87MG was purchased from the American Type Culture Collection (ATCC, Manassas, VA, USA) and was cultured under the conditions recommended by ATCC. The method for preparing T and NK cells is described in the Supplementary Methods.

To detect TNFSF4, TNFRSF18, and IL12RB expression, the cells were stained with anti-TNFSF4-PE (#318706), anti-TNFRSF18-PE (#311603), or anti-IL12RB2-PE (#394205) (BioLegend, San Diego, CA, USA) in the dark at 4 °C for 20 min. Stained cells were analyzed using a CytoFLEX flow cytometer (Beckman Coulter, B rea, CA, USA), and data were analyzed using FlowJo software version 10.1 (Treestar Inc., Ashland, OG, USA).

For Transwell migration assays, 5 × 10^5^ U87MG cells alone or with 5 × 10^5^ T cells were seeded into the lower chamber of a Transwell insert (SPL life science, Pocheon, Korea) in culture medium. After the NK cells were stained with carboxyfluorescein succinimidyl ester (CFSE) (MA 02451, Thermo Fisher Scientific, Waltham, USA), 5 × 10^5^ NK cells were placed to the upper chamber in serum free-medium, and the plates were incubated for 8 h at 37°C. To block TNFSF4, TNFRSF18, and IL-12RB2 expression in T cells, T cells were pre-incubated with 20 μg/mL of the anti-TNFSF4 (MAB10541-SP), anti-TNFRSF18 (MAB689-SP), and anti-IL12RB2 antibodies (AF1959) (R&D Systems, Minneapolis, MN, USA) for 1 hour at 37°C. The number of NK cells that migrated to the lower chamber was determined by automated counting of CFSE-positive cells using a Luna cell counter. Data are presented as the percentage of migration based on total cell input.

## Results

### Clinical information of immune-cell therapy participants

Twelve patients participated in this study based on inclusion and exclusion criteria [10] and received AKC treatment, as summarized in Table 1. Of the 12 patients, five showed durable responses and survived for at least 28 months and for a maximum of 76 months after tumor recurrence; these patients were designated as the responder group. The median OS and PFS times of the responder group were 22.5 and 10 months, respectively. Five patients were alive over 2 years later and were active in their daily lives. The survival time of the seven patients in the non-responder group was less than 2 years, and MRI revealed progression during AKC treatment. The isocitrate dehydrogenase (*IDH1*) mutation (R132H) was detected in only 1 patient in the non-responder group. *MGMT* methylation was found in 3 and 4 patients in the responder and non-responder groups, respectively.

**Table 1.**
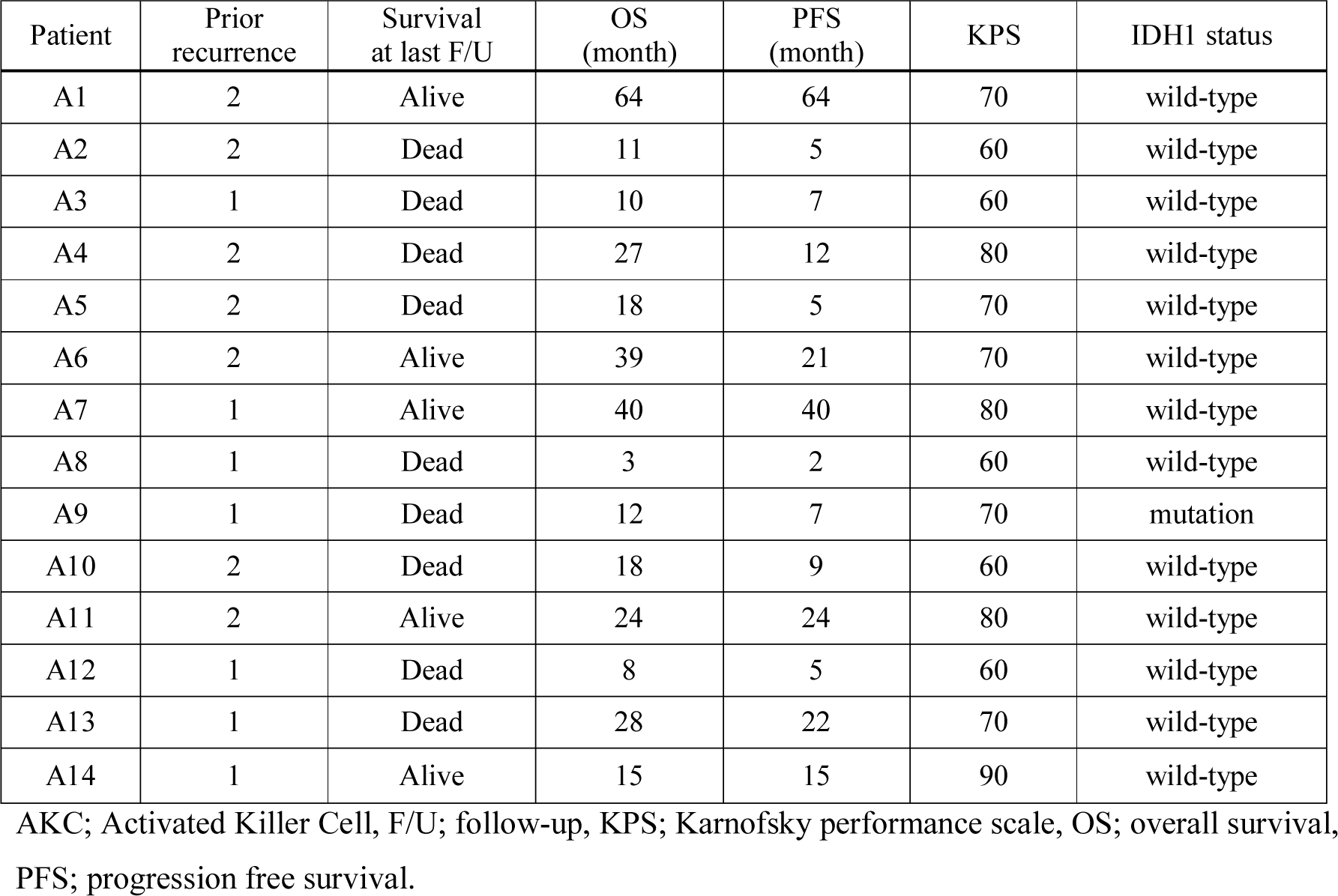
Summary of Patients with GBM who Received AKC immunotherapy.

| Patient | Prior recurrence | Survival at last F/U | OS (month) | PFS (month) | KPS | IDH1 status |
| --- | --- | --- | --- | --- | --- | --- |
| A1 | 2 | Alive | 64 | 64 | 70 | wild-type |
| A2 | 2 | Dead | 11 | 5 | 60 | wild-type |
| A3 | 1 | Dead | 10 | 7 | 60 | wild-type |
| A4 | 2 | Dead | 27 | 12 | 80 | wild-type |
| A5 | 2 | Dead | 18 | 5 | 70 | wild-type |
| A6 | 2 | Alive | 39 | 21 | 70 | wild-type |
| A7 | 1 | Alive | 40 | 40 | 80 | wild-type |
| A8 | 1 | Dead | 3 | 2 | 60 | wild-type |
| A9 | 1 | Dead | 12 | 7 | 70 | mutation |
| A10 | 2 | Dead | 18 | 9 | 60 | wild-type |
| A11 | 2 | Alive | 24 | 24 | 80 | wild-type |
| A12 | 1 | Dead | 8 | 5 | 60 | wild-type |
| A13 | 1 | Dead | 28 | 22 | 70 | wild-type |
| A14 | 1 | Alive | 15 | 15 | 90 | wild-type |

AKC; Activated Killer Cell, F/U; follow-up, KPS; Karnofsky performance scale, OS; overall survival, PFS; progression free survival.

### Immune landscape profiling by the nCounter PanCancer Immune Profiling Panel

To predict the response to AKC treatment, we explored the immune landscape of 12 patients using the nCounter PanCancer Immune Profiling Panel of 730 immune- and cancer-related genes (Figure 1A). We aligned the results of the 730 genes according to functional annotation groups. In Figure 1A, individual patients are represented in each column and are grouped by treatment response: responders (green) on the left and non-responders (red) on the right. Responders showed higher immune-related gene expression compared with non-responders, which suggests a pre-existing increased immune response in the tumor microenvironment of responders. We also calculated the gene signature score of genes in each functional category (Figure 1B). Responders showed high signature scores in the TNF superfamily, cytokines, chemokines, and interleukins, while non-responders showed high signature scores in senescence and cell cycle (AUC ≥ 0.8, Supplementary Table 2).

**Figure 1.**
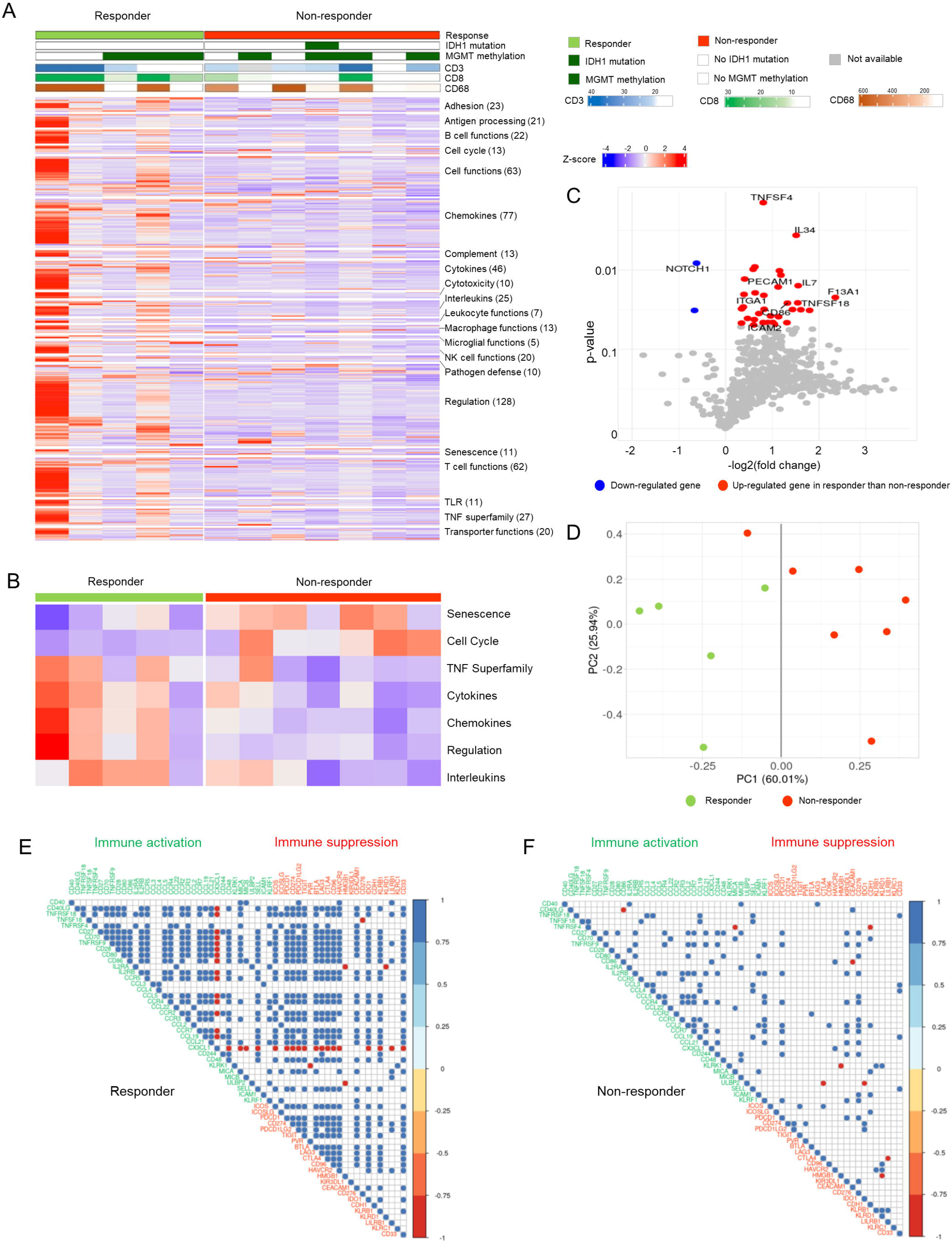
The immune landscape profiling of 12 patients with recurrent GBM. (A) Heatmap of the 730-gene pancancer immune panel for 12 patients. Columns represent patients and rows represent genes. Expression levels were aligned according to the functional annotation groups. Categories of response (responders, non-responders), *IDH1* mutation status, and *MGMT* methylation status are shown. Immunohistochemical stains for CD3, CD8, and CD68 are shown as continuous variables. The mRNA expression levels of genes are represented as colors from red (upregulated, z-score 4) to blue (downregulated, z-score -4). (B) Heatmap of the seven-gene signature scores of the functional annotation groups between responders and non-responders. (C) Volcano plot depicting 35 DEGs between the treatment response groups. (D) A PCA plot based on 35 DEGs. As seen by the dark gray line (PC1 score = 0.00), all patients except one were grouped separately by treatment response. Correlation plots of immune-cell receptors and their ligands in responders (E) and non-responders (F). Pearson’s correlation coefficient scores are represented as colors from blue (1.0) to red (−1.0).

We identified 35 differentially expressed genes (DEGs) based on t-tests where *p* < 0.05; DEGs between responders and non-responders included *TNFSF4* (fold change: 1.75), *IL34* (fold change: 2.85), *IL7* (fold change: 2.94), and *NOTCH1* (fold change: 0.65) (Figure 1C, Supplementary Table 3). Most DEGs (33 DEGs) were highly expressed in responders, but interestingly, *NOTCH1* was highly expressed in non-responders compared with responders. In the principal component analysis (PCA) of 35 DEGs, responders and non-responders, except one patient, were grouped separately by PC1 (Figure 1D). When we compared the expression level of ligands of various NK-activating receptors, such as *MICA, MICB*, and *ULBP2, ULBP2* was significantly overexpressed in responders compared with non-responders (1.52-fold higher, *p* < 0.05, Supplementary Figure 1).

When we analyzed the correlation patterns between immune-cell receptors and their ligands, immune-cell markers were highly correlated in responders (Figure 1E) but not in non-responders (Figure 1F). For example, *TNFRSF18* was highly correlated with 17 immune activating genes, such as *CD40LG, TNFSF4, CD70, TNFRSF9, CD28, CD80, IL2RA, IL2RB, CCR5, CCL5, CCR4, CCR2, CCR3, CCR7, CCL21, MICA*, and *KLRF1* (Pearson’s correlation coefficient: 0.884–0.995 and *p* < 0.05) and 13 immune suppressive genes, such as *ICOS, ICOSLG, PDCD1, CD274, PDCD1LG2, TIGIT, BTLA, CTLA4, CD96, HAVCR2, CEACAM1, IDO1*, and *LILRB1* (Pearson’s correlation coefficient: 0.885–0.960 and *p* < 0.05) in responders. In contrast, only *CCL3, CCL4, CCL19, SELL*, and *CD33* were correlated in non-responders. Interestingly, *CX3CL1* was inversely correlated with most other genes, except *CD244*, in the responder group.

### Identification of genes significantly associated with NK treatment response

To identify genes that are significantly associated with treatment response, we performed a three-step analysis (Figure 2A), and finally, we revealed that *TNFRSF18, TNFSF4*, and *IL12RB2* were candidate biomarkers for predicting the response to AKC treatment. First, we calculated the AUC of each gene and selected 114 genes with AUC values equal to or higher than 0.85. Second, we calculated *p*-values for OS and PFS of each gene and selected 64 genes for which the OS or PFS *p*-value was less than 0.05 (Supplementary Table 4). Figure 2B shows the expression patterns of those 64 genes. Each gene was grouped according to its functional annotation group. Similar to the DEG results, all genes except *NOTCH1* were significantly higher in responders than in non-responders. Lastly, using the 64 genes, we constructed 16 random forest models. One model was constructed using all 64 genes, and the other 15 models were constructed using each functional group or pathway, where the 64 genes were annotated (Supplementary Table 5).9537

**Figure 2.**
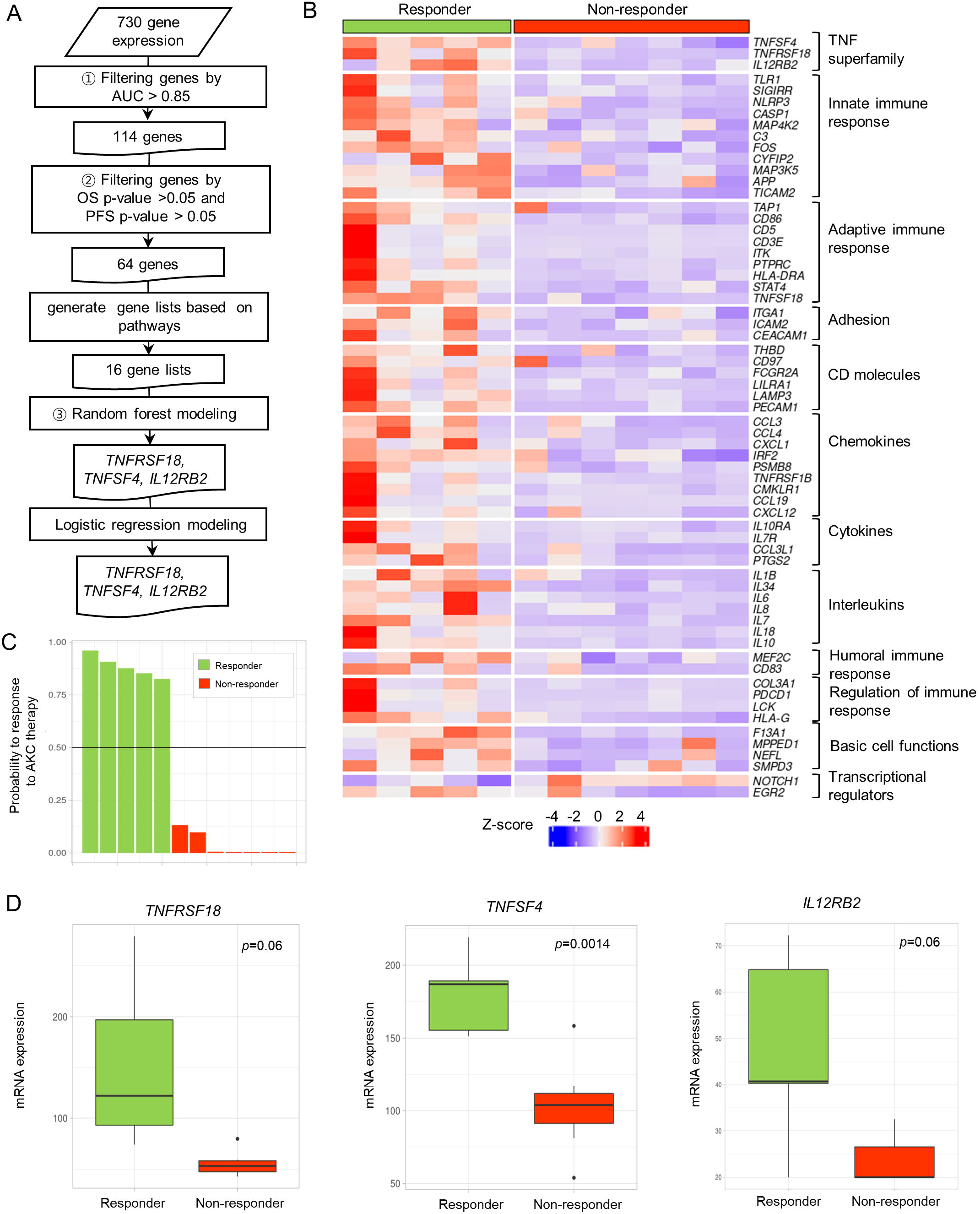
Identification of predictive markers and their expression patterns. (A) Schematic workflow explaining the three steps needed to identify the *TNFSF18, TNFSF4*, and *IL12RB2* genes. (B) Heatmap of 64 genes based on an AUC > 0.85, an overall survival *p*-value > 0.05, and a progression-free survival *p*-value > 0.05. (C) A barplot of probability of response to AKC therapy in a random forest model. According to the black line of probability = 0.5, responders and non-responders were clearly separated. (D) The mRNA expression levels of *TNFRSF18, TNFSF4*, and *IL12RB2* in responders and non-responders are shown as boxplots.

Among the 16 random forest models, the TNF superfamily model of *TNFRSF18, TNFSF4*, and *IL12RB2* showed the best performance with an out-of-bag error rate of 0.0%, a sensitivity of 1.0, a specificity of 1.0, and an accuracy of 100% (95% CI: 73.5%–100%), (Supplementary Table 5). When we calculated the probability of treatment response, the random forest model of these three genes could explicitly separate responders from non-responders (Figure 2C). The same accuracy was achieved when tested using the logistic regression model, which had a sensitivity of 1.0, a specificity of 1.0, and an accuracy of 100% (95% CI: 73.5%–100%). When we compared the expression of each of the three genes according to patient response (Figure 2D), the mRNA expression of the three genes clearly differed according to patients’ treatment response. *TNFRSF18* expression in responders was 2.8-fold higher (*p* = 0.06), that of *TNFSF4* was 1.7-fold higher (*p* = 0.0014), and that of *IL12RB2* was 2.0-fold higher (*p* = 0.06) in responders than in non-responders.

### Validation of qRT-PCR and logistic regression modeling

To validate whether the mRNA expression levels of the three selected genes, *TNFRSF18, TNFSF4*, and *IL12RB2*, could predict the response to AKC treatment, we performed qRT-PCR with different primers and probes and found that the mRNA levels were positively correlated with the expression levels found by NanoString nCounter with high Pearson’s correlation coefficients (0.419 (*TNFRSF18*), 0.700 (*TNFSF4*), and 0.502 (*IL12RB2*)), as shown in Figure 3A. All three genes showed significant differences in expression between responders and non-responders (*TNFRSF18*: *p* = 0.009, *TNFSF4*: *p* = 0.005, and *IL12RB2*: *p* = 0.034) (Figure 3B). Next, we assessed whether the prediction model could distinguish responders from non-responders using the selected genes. The logistic regression model had high accuracy (100%; 95% CI: 73.54%–100%) when validated using the leave-one-out cross-validation. Notably, qRT-PCR for the above three genes and logistic regression modeling are both highly accurate for identifying responders.

**Figure 3.**
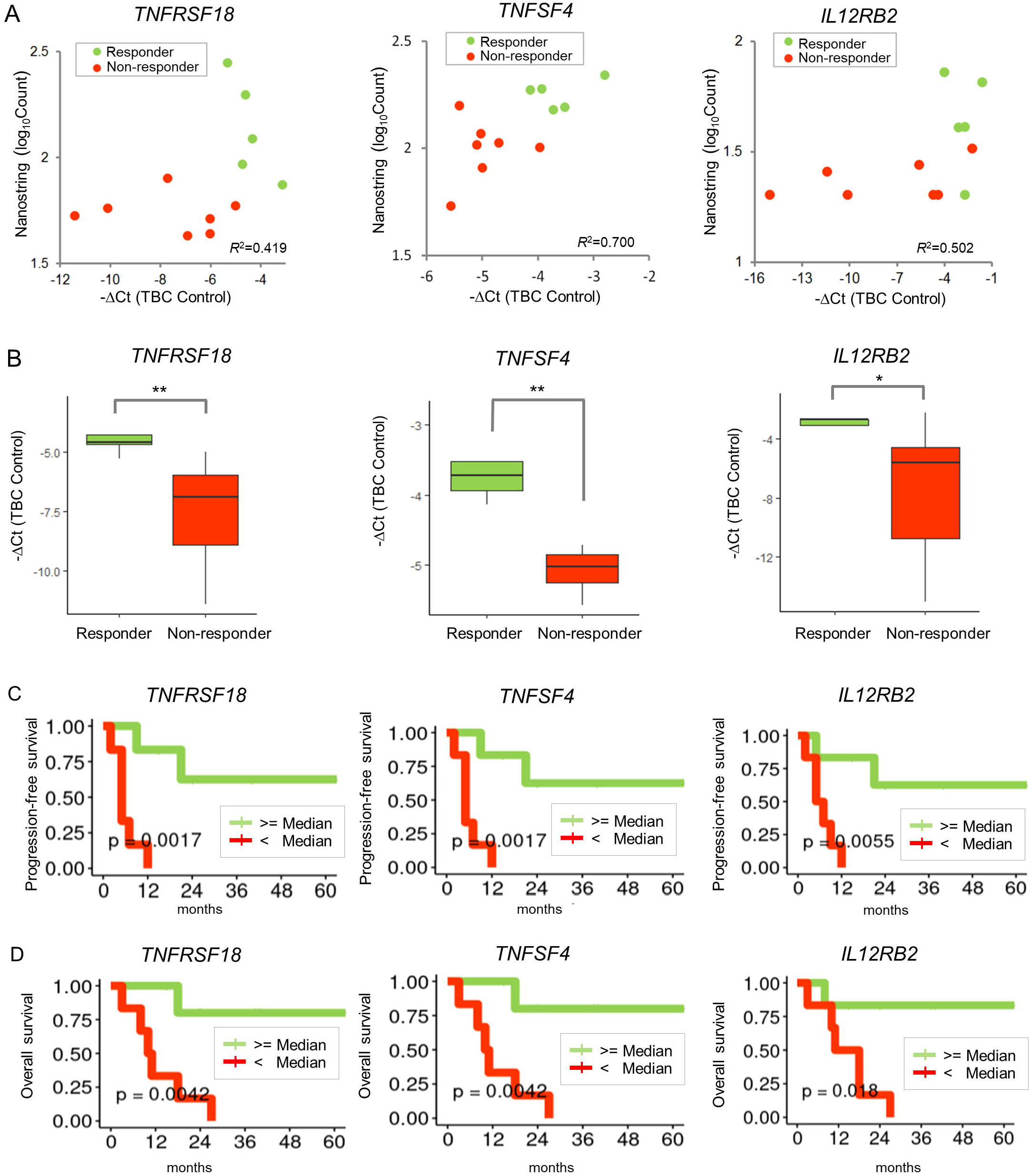
Validation of marker expression and clinical outcomes. (A) Scatter plots of the expression levels by NanoString nCounter and those by qRT-PCR. Patients are represented by different colors according to the response group. Pearson’s correlation coefficients are shown. (B) Boxplots of qRT-PCR quantification for the response group (responders and non-responders). Significance levels of the Wilcoxon rank-sum test are shown as \**p* < 0.05, and \*\**p* < 0.01. (C) Progression-free survival curves according to expression of the three genes. (D) Overall survival curves.

### Clinical implication of the three candidate genes

We investigated the PFS and OS associated with expression of these three genes in enrolled patients. After dividing patients into two groups (high expression and low expression groups) based on the median expression level of each gene, we investigated whether the prognosis of patients in the two groups was associated with expression of these genes. In terms of PFS (Figure 3C), all three genes were significantly associated with patient prognosis (*TNFRSF18*: *p* = 0.0017, *TNFSF4*: *p* = 0.0017, and *IL12RB2*: *p* = 0.0055). In terms of OS (Figure 3D), all three genes were also statistically significantly associated with patient prognosis (*TNFRSF18*: *p* = 0.0042, *TNFSF4*: *p* = 0.0042, and *IL12RB2*: *p* = 0.018). For both PFS and OS, patients with higher expression of all three genes exhibited a better prognosis than those with lower expression.

### The expression of immune-cell markers by immunohistochemistry

To explore the composition of the immune cells in the tumor and peritumoral areas, the T cell marker CD3, the cytotoxic T cell marker CD8, and the macrophage/microglia marker CD68 were assessed in responders and non-responders (Supplementary Table 6). To determine the impact of infiltration of these immune cells on the response to AKC treatment and to adjust for variation in immune-cell infiltration between areas, the mean density per mm^2^ was calculated in the tumor and peritumoral areas of each sample. The median numbers of infiltrating CD3^+^ T cells per mm^2^ were not different between responders and non-responders (data not shown). The mean number of infiltrating cytotoxic CD8^+^ T cells per mm^2^ in the tumor area in responders was 11.5-fold higher than in non-responders (169.6 ± 306.8 vs. 14.62 ± 12.2) (Figure 4A). Although not statistically significant (*p* = 0.32) because of the small number of samples, higher numbers of CD8^+^ T cells per mm^2^ tended to predict AKC treatment response with a high AUC of 0.91 (Figure 4B). A higher CD8^+^ T cell count could discriminate the treatment response with a sensitivity of 0.8, a specificity of 0.86, and an accuracy of 0.83 at a cut-off score of 19.225. For the non-tumor area, the mean number of infiltrating CD8^+^ T cells per mm^2^ in responders was 5.1-fold higher in the tumor area than in non-responders (82.7 ± 170.5 vs. 16.1 ± 17.3); however, this difference was not statistically significant (*p* = 0.43) (Figure 4A).

**Figure 4.**
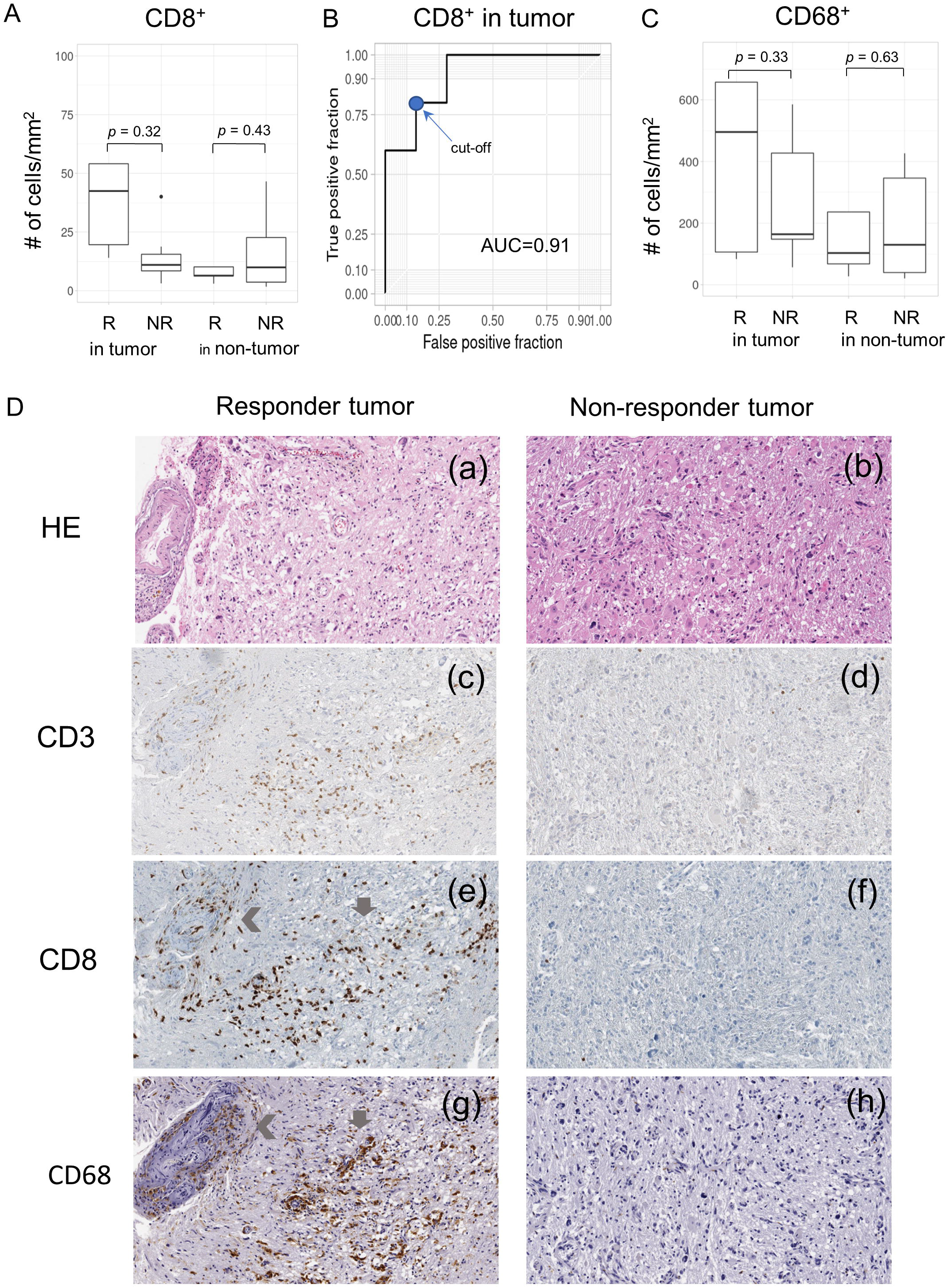
The expression of immune-cell markers (CD8^+^ and CD68^+^ cells) by immunohistochemistry. (A) Distribution of cells expressing the cytotoxic T cell marker CD8^+^ per mm^2^ in tumor and non-tumor areas. R represents responders and NR represents non-responders. (B) The ROC curve based on CD8^+^ cells in the tumor area of each patient. The cut-off score for calculating sensitivity and specificity is indicated by a blue dot. (C) Distribution of cells expressing the macrophage/microglia marker CD68^+^ per mm^2^ in tumor and non-tumor areas. (D) (a-b): Histologic features of the tumor area in responders and non-responders (Hematoxylin and eosin. 1000 × 1000 μm), (c-d) The median number of CD3^+^ T cells was not significantly different between responders and non-responders, as a similar density of CD3^+^ T cell infiltration was observed in both groups. (e-f) The density (number of cells/mm^2^) of CD8^+^ cytotoxic T cells in the tumor was higher in responders than in non-responders. (g-h) The density of CD68^+^ macrophages/microglia in the tumor was higher in responders than in non-responders, and in some areas in responders, increased density of these cells coincided with high CD8^+^ T cell density, which suggests intimate interactions with cytotoxic T cells (grey arrows). In some thick-walled vessels, a high density of immune-cell infiltration was observed (left angle bracket).

The mean number of infiltrating CD68^+^ macrophages/microglia per mm^2^ in the tumor area in responders was 2.2-fold higher than in non-responders (621.2 ± 684.7 vs. 279.5 ± 198.1), but this difference was not statistically significant (*p* = 0.33). The number of these cells in the non-tumor area was not significantly different between responders and non-responders (295.0 ± 424.1 vs. 189.2 ± 184.3, *p* = 0.62) (Figure 4C). The numbers of CD8^+^ and CD68^+^ cells were highly correlated with each other in responders (Pearson’s correlation coefficient = 0.95, *p* = 0.013, Supplementary Figure 2A), which suggests that these immune cells might influence each other’s infiltration in the in responders (Figure 4D), whereas it was not in non-responders.

Based on this result, to examine the pattern of distribution of CD8^+^ cytotoxic T cells and CD68^+^ macrophages/microglia in the tumor microenvironment of responders and non-responders, a CD8^+^ and CD68^+^ cell density map was generated (See Supplementary Methods). In responders, the high-density areas of CD8^+^ and CD68^+^ cells seemed to be located more frequently in tumor tissue rather than in non-tumor tissue, and similar distribution patterns were observed within the same piece of tissue. This suggests that cases with close interaction of CD8^+^ and CD68^+^ cells in the tumor might respond better to NK cell therapy. In contrast, non-responders showed somewhat nonspecific distribution patterns of high-density CD8^+^ and CD68^+^ cell areas (Supplementary Figure 2B). The infiltration of these two immune cells using immunohistochemical stain was correlated with mRNA expression of each genes by NanoString in the responders, whereas their correlation was not seen in non-responders (Supplementary Figure 2C).

In addition, we found that infiltration by CD8^+^ cytotoxic T cells was highly correlated with *TNFRSF18* and *TNFSF4* expression (Pearson’s correlation coefficient = 0.83, *p* = 9.0 × 10^4^ for *TNFRSF18*, Pearson’s correlation coefficient = 0.56, *p* = 0.06 for *TNFSF4*) (Supplementary Figure 3C).

### Effect of three candidate genes on NK cell migration

To validate whether expression of the three candidate genes (*TNFSF4, TNFRSF18*, and *IL12RB2*) in the tumor microenvironment is implicated in NK cell migration toward cancer cells, we conducted a Transwell migration assay using GBM U87MG cells, T cells, and NK cells derived from PBMCs. First, we evaluated the expression of these three candidate genes on T cells and U87MG cells. The three candidate genes were highly expressed on activated T cells (62.29%, 63.49%, and 69.59% for *TNFSF4, TNFRSF18*, and *IL12RB2*, respectively) (Figure 5A).

**Figure 5.**
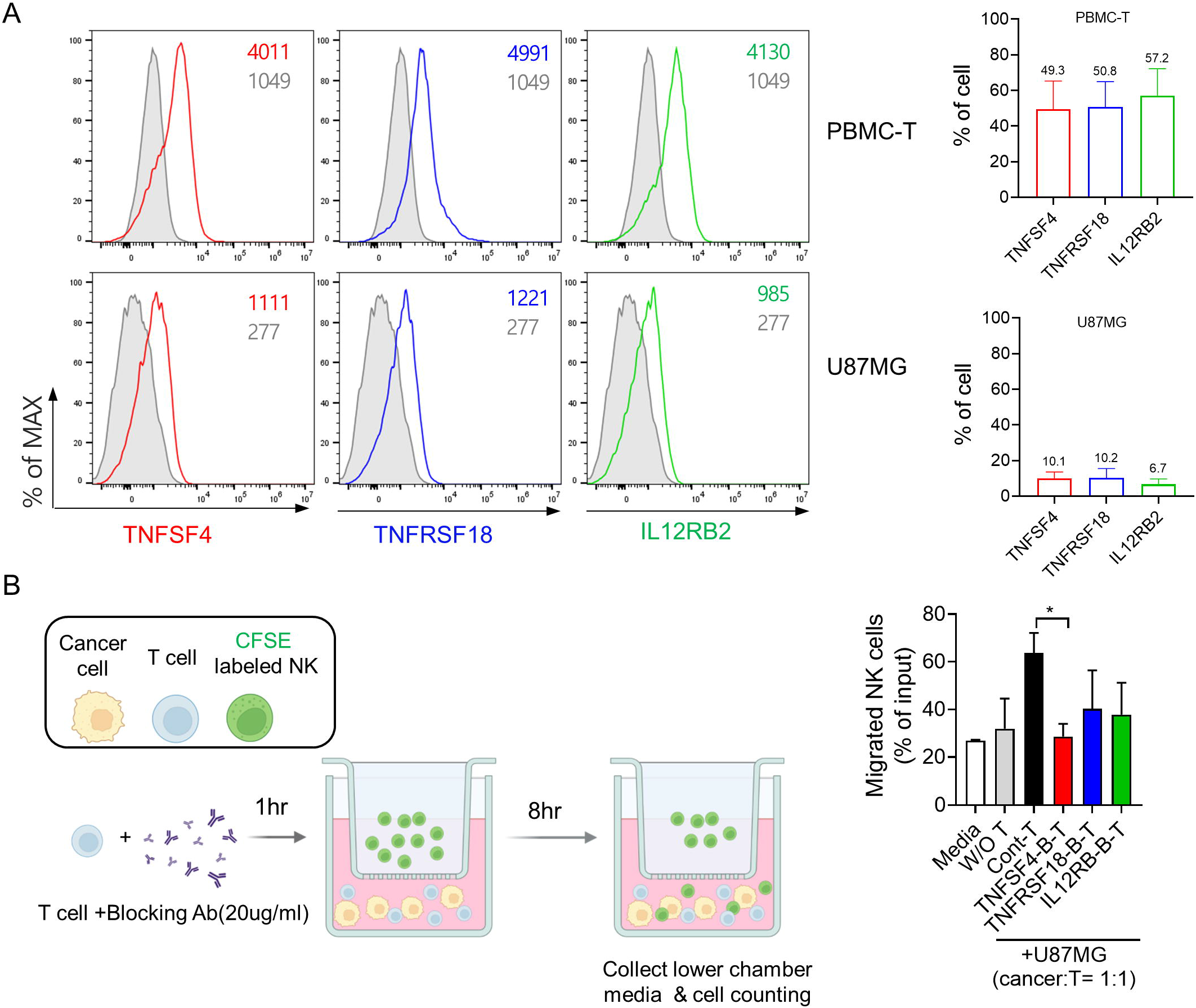
Effects of three candidate genes on migration of NK cells toward GBM cancer cells. (A) Expression of TNFSF4, TNFRSF18, and IL12RB2 in T cells and U87MG cells was analyzed by flow cytometry. Left: Representative histogram plots. Gray histograms represent isotype controls. Right: Column graphs show the frequencies of TNFSF4^+^, TNFRSF18^+^, and IL12RB2^+^ cells. (B) Transwell migration assay: CFSE-stained NK cells were plated in the upper chamber of a Transwell insert with an 8-mm pore size. Cells were allowed to migrate for 8 hours toward U87MG cells with or without T cells in the lower chamber. T cells were pre-incubated with blocking antibodies to TNFSF4, TNFRSF18, and IL12RB2 for 1 hour. NK cells that migrated to the lower chamber were counted using a cell counter. The experiments were performed in triplicate.

After blocking expression of these three candidate genes on T cells, we compared the percentage of NK cells that migrated from the upper chamber to the lower chamber between the GBM + control T cell group and the GBM + T cell groups in which expression of these genes was blocked. The number of NK cells that migrated toward U87MG GBM cells was significantly reduced in the *TNFSF4*-blocked T cell group (26.9% ± 2.0%, *p* = 0.027), and tended to decrease in the *TNFRSF18*-blocked T cell group (42.1% ± 9.6%, *p* = 0.195) and the *IL12RB*-blocked T cell group (37.8% ± 9.6%, *p* = 0.146) compared with the control T cells in which the expression of these three genes was not blocked (63.8% ± 5.9%) (Figure 5B). Collectively, these results suggest that higher expression of these three genes, especially *TNFSF4*, in T cells contributed to NK cell migration toward cancer cells.

## Discussion

GBM is the most common and aggressive primary malignant brain tumor and patients with GBM have a very poor prognosis, as their median OS is 14.6 months [16]. Although newly developed immuno-oncology drugs have been used to treat primary GBM, the current therapeutic approaches have very limited impact on improvement of the survival of patients with GBM. The CheckMate-143 study [4], a large-scale phase 3 study conducted in patients with recurrent GBM, compared Nivolumab, a PD-1 immune checkpoint inhibitor, and Bevacizumab, a control drug, but failed to demonstrate an effect on OS. Other immune checkpoint inhibitors have also not shown any significant therapeutic effects.

Considering the results of research conducted thus far, both the GBM cancer cells themselves and the tumor microenvironment seem to be the cause of the failure of these immuno-oncologic drugs. Since GBM has a low tumor mutation burden compared with other cancers, it has fewer tumor antigens, and therefore, it is believed to be difficult to prime an anticancer immune response in GBM. In addition, GBM cells secrete many soluble immunosuppressive factors, such as TGF-β, prostaglandin E2 (PGE2), vascular endothelial growth factor (VEGF), IL10, CCL2, and CCL22, which suppress cytotoxic T cells and recruit Treg cells. Treg cells contribute to cytotoxic T cell suppression and tumor progression via production of IL10, IL35, and TGF-β. GBM cells also recruit TAMs by releasing CX3CL1 or CCL5, and they promote the acquisition of an M2 phenotype by microglial cells via secretion of IL10, IL4, IL13, and TGF-β. Therefore, NK cell-based immunotherapy might become an attractive and promising therapy that can overcome the immunosuppressive microenvironment of GBM, since NK cells, complement the antitumor activity of T cells by releasing various cytokines or chemokines, by recruiting dendritic cells, and by directly killing cancer cells.

Recently, we performed a clinical trial using intravenous autologous AKC treatment for patients with recurrent GBM after surgical resection to overcome the immunosuppressive TME. In our clinical trial, we observed that this NK therapy suppressed the tumors and resulted in prolonged OS and PFS in patients with some subsets of these tumors [10]. Therefore, we examined the immune profiles in tumor and TME tissues of these patients and compared them between responders and non-responders to identify the biomarkers predictable to treatment responsiveness.

In the present study, we found that the expression of genes related to various immune-signaling molecules was increased in responders. Interestingly, the DEG analysis and filtering of genes by AUC, OS, or PFS revealed that the genes commonly related to immune-cell function, regulation, and recruitment were significantly increased in responders compared with non-responders. When the correlation patterns between immune-cell receptors and their ligands were analyzed, signaling proteins were highly correlated in responders whereas they were not correlated in non-responders (Figure 1E). This immune profiling might be related to higher infiltration of CD8^+^ T cells in responders compared with non-responders, although the difference between the two was not statistically significant, and it was not clear whether these T cells were activated or exhausted.

Considering that previous studies reported the frequent occurrence of transition to a mesenchymal subtype with increased expression of various immune-related genes in recurrent glioblastomas after standard therapy [17, 18], the responder group is more likely to have the mesenchymal molecular subtype, which is considered to be more aggressive. In the mesenchymal subtype of GBM, an increased subpopulation of TAMs/microglia has been described as an integral component that contributes to mesenchymal transition [19, 20]. In this study, we also demonstrated that responders had a 2.2-fold higher infiltration of TAMs/microglia than non-responders, which supports the concept that the responder group was more likely to have the mesenchymal subtype of GBM. In addition, using density maps and correlation analysis by immunohistochemistry, we demonstrated similar distribution patterns and highly correlated infiltration of CD8^+^ and CD68^+^ cells in the tumors of responders. Taken together, our results implied that AKC treatment might be effective at least in some subset of tumors in the highly immune-active group of GBM, such as those with the mesenchymal subtype, which is known to have poor outcomes.

In addition, the DEG analysis demonstrated that *NOTCH1* expression was significantly lower in the responder group than in the non-responder group, whereas the other significantly altered genes were upregulated in the responder group. Considering that NOTCH1 is a marker of glioma stem cells, it is suggested that non-responders harbor more glioma stem cells than responders. When the correlation patterns between immune-cell receptors and their ligands were analyzed, signaling proteins except CX3CL1 were highly and positively correlated in responders, whereas they were not correlated in non-responders (Figure 1E). CX3CL1 is reported to be mostly expressed in neurons where it inhibits the overexpression of pro-inflammatory molecules. Therefore, it is believed that CX3CL1 expression was inversely correlated with the expression of other inflammatory genes in the responder group.

In this study, we identified three candidate genes, *TNFRSF18, TNFSF4*, and *IL12RB2*, which can best predict responsiveness to AKC treatment; these were selected through the random forest method (Figure 3C). These genes are members of the TNF family, which is known to coordinate co-stimulation or co-inhibition of the immune response in the tumor microenvironment.

TNFRSF18, also known as GITR (glucocorticoid-induced tumor necrosis factor receptor related protein), was initially known to play a co-stimulatory role in T cell activation. However, GITR-GITRL interactions are far more complex. GITR is expressed at high levels on T regulatory cells [21] as well as on activated CD4^+^ and CD8^+^ T cells [22] and at intermediate levels on NK cells. According to previous reports, GITR expression on tumor-infiltrating Tregs is higher than that on tumor CD8^+^ T cells and peripheral Treg cells [23]. On the contrary, agonistic GITR antibodies directly suppressed Tregs and reduced the sensitivity of effector T cells to Treg-mediated suppression [24]. A current clinical trial reported that an agonistic anti-GITR mAb reduced Treg levels both in the circulation and the tumors of treated patients [25, 26]. Considering the previous reports and the finding that our NK cells expressed high GITR ligands (2.65-fold higher than PBMCs, data not shown), GITR-GITRL signaling might have induced anticancer immunity by Treg suppression when we introduced our NK therapeutics.

TNFSF4, also known as OX40 ligand, is primarily expressed on antigen-presenting cells, activated T cells, and some endothelial cells, where it stimulates OX40 expression on the surface of activated T or NK cells. OX40-OX40L signaling increases the adaptive immune response to pathogenic antigens by promoting effector and memory T cell survival [27]. The previous study using The Cancer Genome Atlas and the Cancer Cell Line Encyclopedia RNA sequencing data from anti-PD1-treated patients with melanoma demonstrated that the low *TNFSF4* mRNA expression group was associated with worse prognosis and showed a worse outcome after anti-PD1 treatment [28]. On the contrary, a recent analysis [29] of a TNF family based-signature in diffuse gliomas using 4 datasets reported that 35 TNF family genes including *TNFSF4* were related to the high-risk group of patients, who had a poor prognosis. However, several preclinical studies using a murine glioma model [30, 31] demonstrated that OX40-OX40L activation was related to a better prognosis and prolonged survival by agonistic OX40 antibody therapy through reverse intracranial T cell exhaustion. The findings from previous preclinical research [32] that OX40 on NK cells interacts with OX40L on dendritic cells resulting in increased IFN-γ release and T cell induction, and from our previous report that shows a high-level of OX40 in our AKC cells [10, 33], provide reasonable evidence for OX40L as a predictive biomarker of our AKC therapeutics. According to our Transwell migration assay, it is assumed that T cells expressing TNFSF4 in the TME of GBM might induce NK cell migration into the tumor, resulting in a better therapeutic effect of our AKC treatment.

*IL12RB2* encodes the β2 chain of the high-affinity receptor that binds to IL-12, which is well-known cytokine that bridges innate and adaptive immunity [34]; the β2 chain is expressed by antigen-presenting cells, including macrophages and microglia. IL-12 drives T helper responses, enhances T and NK cell cytotoxicity, and induces IFN-γ production by T and NK cells [35]. IL12RB2 is essential for IL-12 signal transduction and functions as a tumor suppressor. Higher expression of *IL12RB2* was reported to be associated with better prognosis and longer survival in various human tumors, including lung and laryngeal cancers [36, 37]. Taken together, it could be expected that higher expression of *IL12RB2* is predictive marker for responsiveness to NK cell therapeutics.

In conclusions, we identified *TNFRSF18, TNFSF4*, and *IL12RB2* as biomarkers that predict response to NK cell therapeutics by studying the tumor immune microenvironment using NanoString and qRT-PCR analyses in patients with recurrent GBM who received activated NK cell treatment. Given the fact that the early clinical trial sample size is usually small, however, those samples are worthy for predicting clinical efficacy, the results would be very valuable although the sample size was small in this study. Therefore, these three genes can be used to screen individuals who might benefit from NK cell therapy. In addition, our results show that responders to NK therapy have an immune-activated TME, whereas non-responders do not. Considering that immune and inflammatory response-enrichment is related to the high-risk group in this tumor type, our findings highlight a new treatment strategy of NK cell therapeutics for patients with highly aggressive recurrent GBM.\

## Data Availability

All expression data are available in the GEO Database (https://www.ncbi.nlm.nih.gov/geo/) under accession number GSE207753.

## List of abbreviations

AKC: Autologous activated NK cells
ATCC: American Type Culture Collection
AUC: Area under the ROC curve
CFSE: Carboxyfluorescein succinimidyl ester
CI: Confidence interval
DEG: Differentially expressed genes
FFPE: Formalin-fixed paraffin-embedded
GBM: Glioblastoma multiforme
KHIDI: Korea Health Industry Development Institute
NK: Natural killer
OS: Overall survival
PBMC: Peripheral blood mononuclear cells
PCA: Principal component analysis
PFS: Progression-free survival
TAM: Tumor-associated macrophages
TMB: Tumor mutation burden
Treg: Regulatory T

## Declarations

### Ethics approval and consent to participate

This study was approved by the Institutional Review Board of Bundang CHA Medical Center (#2012-12-172, #2021-01-024). Informed consent was obtained from each patient before the clinical trial.

### Consent for publication

The authors approved the manuscript and gave their consent for submission and publication.

### Author Contributions

Conceptualization, H.K., H.J.A., J.L. K.C. and S.H.; methodology, H.K., H.J.A., J.L. and S.H.; software, J.G.J. and S.H.; validation, D.J., J.G.J. and J.Y.J.; visualization, D.J., H.K., J.G.J. and S.H.; formal analysis, J.G.J. and S.H.; investigation, J.Y.J. and D.J.; resources, H.K., J.H., H.J.A., J.L. and S.H.; data curation, H.K. and J.H.; writing □ original draft preparation, J.L., H.J.A. and S.H.; writing □ review and editing, J.L., H.K., H.J.A. and S.H., supervision, K.J. and H.J.A., project administration, K.J. and H.J.A., funding acquisition, H.J.A. and S.H. All authors have read and agreed to the published version of the manuscript.

