## Supplementary figure for "Predictive Biomarkers for the Responsiveness of Recurrent Glioblastomas to Activated Killer Cell Immunotherapy"

### Supplementary figures

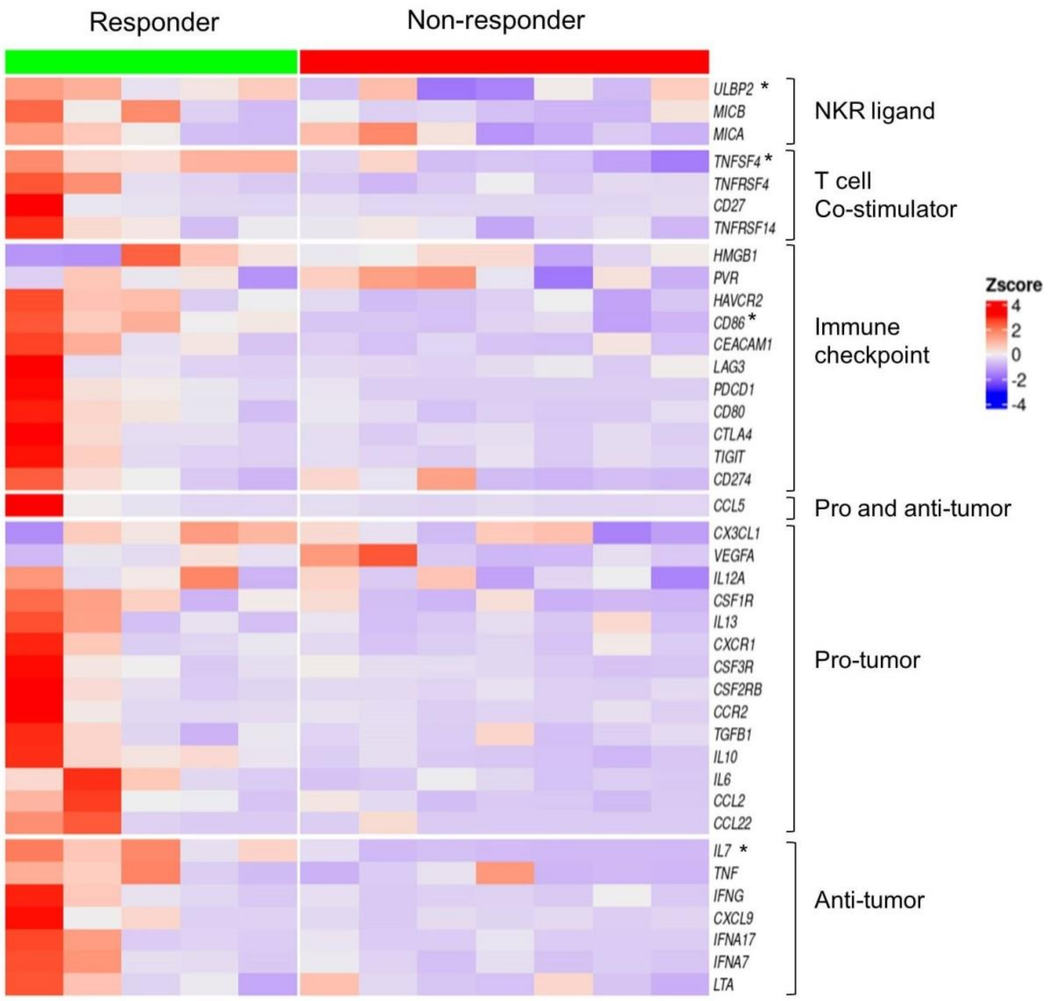


**Supplementary Figure 1. The expression patterns of ligands of various NK-activating receptors.** Heatmap for 40 genes related to immune response including NK-activating receptors from 12 patients. Expression levels were aligned according to functional annotation groups. Patients were grouped by treatment response. Responders are in green and non-responders are in red. Genes with a *p*-value < 0.05 are denoted with an asterisk (*). The mRNA expression levels are represented as colors from red (up-regulated, z-score 4) to blue (down-regulated, z-score −4).

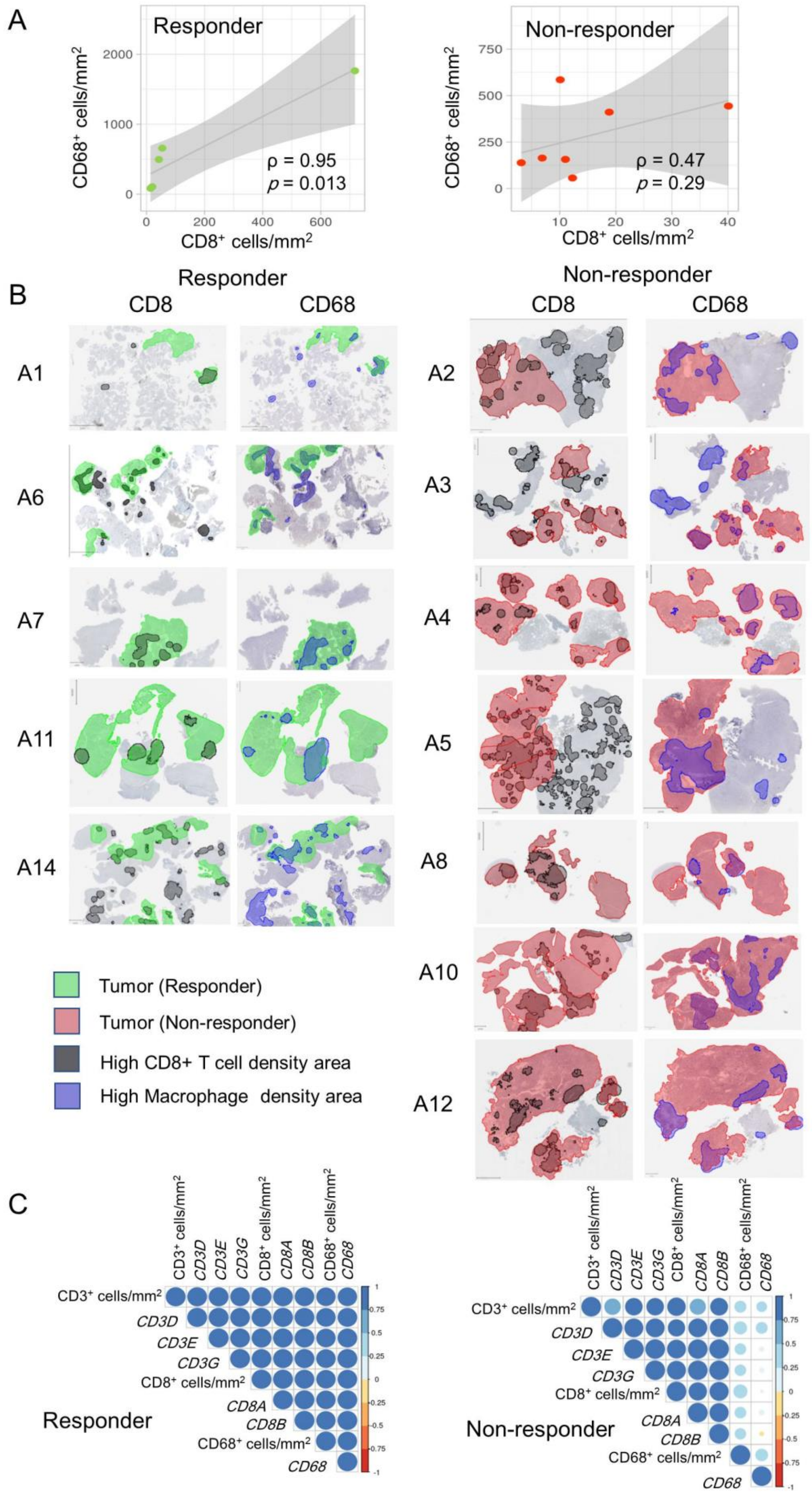


**Supplementary Figure 2. The correlation of CD8$^{+}$ and CD68$^{+}$ cells in the tumor area.** (A) The dot plots between CD8$^+$ cells and CD68$^+$ cells in the tumor area, the trend line, and confidence interval (gray area) between them. In responders, they are significantly correlated (Pearson's correlation coefficient = 0.95, $p$ = 0.013), but in non-responders, they are not (Pearson's correlation coefficient = 0.47, $p$ = 0.29). (B) Density map of high-density areas of CD8$^+$ and CD68$^+$ cells. In responders, the high-density areas of CD8$^+$ and CD68$^+$ cells seemed to be located more frequently in the tumor tissue rather than the non-tumor tissue, with similar distribution patterns within the same piece of tissue. This suggests that the cases with close interaction of CD8$^+$ and CD68$^+$ cells in the tumor might respond better to NK cell therapy. In contrast, non-responders showed somewhat nonspecific distribution patterns of high-density CD8$^+$ and CD68$^+$ cell areas. Only a few areas showed similar distributions in the same piece of tissue from non-responders. (C) The correlation plots between protein expression by immunohistochemistry and the mRNA expression of each gene by NanoString in responders and in non-responders. In responders, they are significantly correlated, but in non-responders, CD68 were not significantly correlated to others.

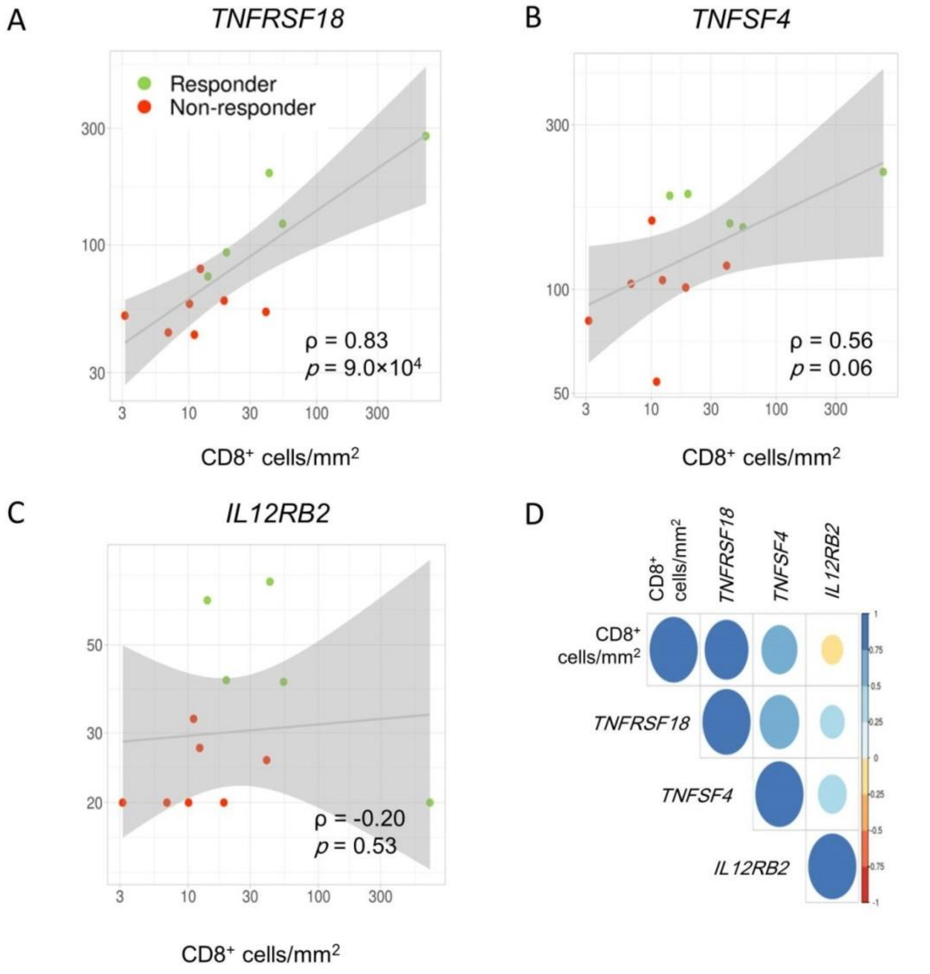


**Supplementary Figure 3. The correlation of CD8$^+$ cytotoxic T cells by immunohistochemical staining and mRNA expression of three genes by Nanostring.** The dot plots show the correlation between CD8$^+$ cells in the tumor area and mRNA expression levels of *TNFRSF18*, *TNFSF4*, and *IL12RB2* according to the NanoString analysis. (A) *TNFRSF18* and was significantly correlated with CD8$^+$ cells in the tumor area (Pearson's correlation coefficient = 0.83, $p$ = 9.0 × $10^4$). (B) *TNFSF4* was also correlated with CD8$^+$ cells in the tumor area (Pearson's correlation coefficient = 0.56, $p$ = 0.06). (C) *IL12RB2* was not correlated with CD8$^+$ cells in the tumor area (Pearson's correlation coefficient = −0.20, $p$ = 0.53). (D) The correlation plot also shows the same pattern between these genes and CD8$^+$ cells. Pearson's correlation coefficient scores are represented as colors from blue (1.0) to red (−1.0). The size of the circle represents the statistical significance; the bigger the circle, the greater the significance.
