## Supplementary methods for "Predictive Biomarkers for the Responsiveness of Recurrent Glioblastomas to Activated Killer Cell Immunotherapy"

### *RNA Extraction*

For RNA extraction, 2-10 sections of 5 μm-thick formalin-fixed paraffin-embedded (FFPE) tissue were prepared after selection of one representative block which contained tumor and tumor microenvironment. The FFPE tissues were deparaffinized and homogenized using the Covaris Focused-ultrasonicator (Covaris, Mannheim, Germany). The truXTRAC FFPE total NA Ultra Kit (Covaris, Mannheim, Germany) was used for efficient and sequential extraction of RNA from FFPE tissue samples using Adaptive Focused Acoustics (AFA) according to manufacturer’s instructions. RNA concentration and integrity were determined using Qubit$^{TM}$ RNA HS Assay Kit with the Qubit$^{TM}$3 Fluorometer (TermoFisher Scientifc, Waltham, MA, USA) and an Agilent TapeStation (Agilent Technologies, Amstelveen, the Netherlands). RNA quality was reflected by RIN (RNA Integrity Number) and DV200 values (% of RNA fragments >200 nucleotides) utilizing the Agilent TapeStation software 3.2.

### *NanoString nCounter Analysis*

The extracted RNAs were run on the NanoString nCounter Analysis System (NanoString Technologies, Inc., WA, USA) according to the manufacturer's instructions. Each RNA (5 μL, 100–300 ng) was mixed with 8 μL of Master Mix (reporter codeSet and hybridization buffer). The capture probeSet (2 μL) of the nCounter PanCancer Immune Profiling Panel was added after which the samples were placed in a 65°C thermocycler (Bio-Rad Laboratories Inc., Hercules, California, USA) for 16 hours. The samples were transferred to the preparation station (NanoString Technologies) with prepared nCounter Master Kit and a cartridge. The preparation station runs for approximately 2.5 to 3 hours. The cartridges were transferred to the Digital Analyzer (NanoString Technologies) and were scanned on the Digital Analyzer at 555 fields of view.

### *Data preprocessing and analysis*

nSolverTM Analysis Software version 4.0 and NanoString QCPro version 1.14.0 were used to perform a quality check of the NanoString nCounter data. For data normalization for quantifying gene expression values, we used geNorm algorithm [1] in nCounter Advanced Analysis version 2.0.115. The nCounter PanCancer Immune Profiling Panel includes 770 genes related to immune response and general cancer processes. The normalized 730-gene expression data, except 40 internal reference housekeeping genes, were analyzed for predictability by AUC (Area Under the ROC Curve) score. To identify the association of each gene to prognosis, we also calculated p-values for OS and PFS. For multiple comparisons, we used the Benjamini-Hochberg procedure. Differentially expressed genes (DEGs) between responders and non-responders were identified by t-test p-value <0.05. The gene signature scores of genes in the 21 functional categories provided by nCounter PanCancer Immune Profiling Panel were measured by single-sample gene set enrichment analysis (ssGSEA) using the GSVA R package [2].

### *The preparation of T and NK cells*

For preparing T and NK cells, human Peripheral blood mononuclear cells (PBMCs) were collected from healthy donors via leukapheresis. All healthy donors provided written informed consent prior to conducting the study. CD3+ T cells and CD3-CD56+ NK cells were obtained through CD3+ or CD3-/CD56+ selection with anti-CD3 (Miltenyi Biotec, Bergisch Gladbach, Germany) and anti-CD56 antibodies (Miltenyi Biotec) using the closed and automated platform, Prodigy (Miltenyi Biotec). Purified CD3+ T cells were activated using Human CD3/CD28/CD2 T Cell Activator (Stem cell technology, Vancouver, Canada) and 100 IU/mL recombinant human IL-2 (Novartis, Basel, Switzerland) and cultured for 4 days. Purified CD3-CD56+ NK cells cultured for 14 days with 1000 IU/mL recombinant human IL-2, 50 ng/mL recombinant human IL-18 (R&D system, Minneapolis, MN, USA), which was the same expansion method with AKC therapeutics [3].

### *Density maps of $CD8^+$ and $CD68^+$ cells*

To examine the distribution patterns of the CD8+ cytotoxic T cells and CD68+ macrophages/microglia in tumor and non-tumor areas of each patient, density maps were generated on QuPath [4]. The areas with CD8+ or CD68+ cell densities higher than the mean number of positive cells/mm2 of the tumor area of each patient were visualized (solid red color). These areas were defined as high-density areas.

### *Statistical analysis*

All plots such as volcano plots and bar plots were depicted in ggplot2 R. AUC score was measured by ROCR R package [5] and PFS p-value was calculated by log rank test in survival R package. Survival analysis was done by survival and survminer R packages. Principal Component Analysis (PCA) was done by prcomp function in stats package. Heatmap analysis was carried out with ComplexHeatmap R package. Correlation heatmaps were depicted in corrplot package. All statistical data were analyzed using R 4.1.0.
