## Supplementary Table for "Predictive Biomarkers for the Responsiveness of Recurrent Glioblastomas to Activated Killer Cell Immunotherapy"

### Supplementary tables

**Supplementary Table 1. Immunohistochemical stain reagents and staining conditions.**

| Antibody | Company | Antigen retrieval time (min) | Antibody application time (min) |
|---|---|---|---|
| CD3(2GV6) | Roche Diagnostics | 36 | 32 |
| CD8(SP57) | Roche Diagnostics | 64 | 32 |
| CD68(KP-1) | Roche Diagnostics | 64 | 32 |

**Supplementary Table 2. Seven signature scores of functional annotation groups discriminating responder and non-responder patients.**

| Function | AUC | t-test p-value | adjusted p-value |
|---|---|---|---|
| Senescence | 0.886 | 0.026 | 0.268 |
| Cell cycle | 0.914 | 0.009 | 0.185 |
| Chemokines | 0.829 | 0.074 | 0.357 |
| Regulation | 0.829 | 0.106 | 0.357 |
| Cytokines | 0.829 | 0.119 | 0.357 |
| TNF superfamily | 0.800 | 0.094 | 0.357 |
| Interleukins | 0.829 | 0.082 | 0.357 |

AUC; Area Under a ROC Curve.

**Supplementary Table 3. 35 differentially expressed genes between responders and non-responders.**

| Gene symbol | AUC | t-test p-value | adjusted p-value | average expression of responder | average expression of non-responders | Fold change | DEG |
|---|---|---|---|---|---|---|---|
| *TNFSF4* | 0.94 | 0.0014 | 0.49 | 180.41 | 103.17 | 1.75 | up |
| *IL34* | 1.00 | 0.0036 | 0.49 | 180.06 | 63.20 | 2.85 | up |
| *NOTCH1* | 0.97 | 0.0082 | 0.49 | 1629.68 | 2508.00 | 0.65 | down |
| *CD47* | 0.91 | 0.0091 | 0.49 | 5496.61 | 3558.92 | 1.54 | up |
| *NFKB1* | 0.94 | 0.0098 | 0.49 | 340.64 | 226.26 | 1.51 | up |
| *HLA-G* | 0.97 | 0.0102 | 0.49 | 2901.93 | 1304.42 | 2.22 | up |
| *FOS* | 0.94 | 0.0116 | 0.49 | 29964.84 | 13190.11 | 2.27 | up |
| *IRF2* | 0.89 | 0.0130 | 0.49 | 1018.54 | 767.44 | 1.33 | up |
| *IL7* | 1.00 | 0.0157 | 0.49 | 65.65 | 22.37 | 2.94 | up |
| *PECAM1* | 1.00 | 0.0164 | 0.49 | 2548.55 | 1168.98 | 2.18 | up |
| *APP* | 0.91 | 0.0194 | 0.49 | 27727.14 | 17885.43 | 1.55 | up |
| *TICAM2* | 0.97 | 0.0203 | 0.49 | 308.30 | 234.56 | 1.31 | up |
| *ITGA1* | 0.94 | 0.0209 | 0.49 | 1221.72 | 693.15 | 1.76 | up |

| | | | | | | | |
|---|---|---|---|---|---|---|---|
| *F13A1* | 1.00 | 0.0221 | 0.49 | 2011.60 | 395.02 | 5.09 | up |
| *TNFSF18* | 0.94 | 0.0259 | 0.49 | 143.85 | 49.31 | 2.92 | up |
| *CD86* | 1.00 | 0.0261 | 0.49 | 390.29 | 156.01 | 2.50 | up |
| *CD58* | 0.83 | 0.0292 | 0.49 | 1011.33 | 777.55 | 1.30 | up |
| *UBC* | 0.86 | 0.0308 | 0.49 | 84150.48 | 66348.47 | 1.27 | up |
| *C1S* | 0.89 | 0.0313 | 0.49 | 2519.39 | 1421.60 | 1.77 | up |
| *C3* | 0.97 | 0.0315 | 0.49 | 13865.90 | 5139.26 | 2.70 | up |
| *STAT4* | 0.94 | 0.0317 | 0.49 | 139.98 | 45.86 | 3.05 | up |
| *CCL3L1* | 0.91 | 0.0323 | 0.49 | 10675.48 | 3070.79 | 3.48 | up |
| *MAVS* | 0.91 | 0.0323 | 0.49 | 1343.46 | 2129.56 | 0.63 | down |
| *MEF2C* | 0.89 | 0.0354 | 0.49 | 3012.08 | 1839.02 | 1.64 | up |
| *EGR2* | 0.86 | 0.0380 | 0.49 | 955.87 | 490.28 | 1.95 | up |
| *CASP1* | 0.91 | 0.0388 | 0.49 | 1024.77 | 466.99 | 2.19 | up |
| *SMPD3* | 0.89 | 0.0408 | 0.49 | 69.50 | 50.14 | 1.39 | up |
| *CYLD* | 0.89 | 0.0422 | 0.49 | 2462.42 | 1607.89 | 1.53 | up |
| *ICAM2* | 0.97 | 0.0458 | 0.49 | 405.57 | 201.50 | 2.01 | up |
| *REL* | 0.89 | 0.0459 | 0.49 | 681.46 | 363.99 | 1.87 | up |
| *MAP3K5* | 0.86 | 0.0459 | 0.49 | 2720.01 | 1558.23 | 1.75 | up |
| *ICAM1* | 0.86 | 0.0461 | 0.49 | 2065.60 | 831.59 | 2.48 | up |
| *MAPK3* | 0.80 | 0.0466 | 0.49 | 2334.33 | 1843.12 | 1.27 | up |
| *CXCL5* | 0.83 | 0.0483 | 0.49 | 95.45 | 46.30 | 2.06 | up |
| *ULBP2* | 0.83 | 0.0494 | 0.49 | 113.94 | 75.15 | 1.52 | up |

AUC; Area Under a ROC Curve, Fold change; responder/non-responder, DEG; up or down in responder compared to non-responder.

**Supplementary Table 4. 64 significantly associated genes to therapy's response.**

| Gene symbol | AUC | fold change | OS p-value | OS adjusted p-value | PFS p-value | PFS adjusted p-value | Functional category |
|---|---|---|---|---|---|---|---|
| *APP* | 0.91 | 1.55 | 0.004 | 0.130 | 0.006 | 0.104 | |
| *C3* | 0.97 | 2.70 | 0.010 | 0.168 | 0.006 | 0.104 | Regulation |
| *CASP1* | 0.91 | 2.19 | 0.004 | 0.130 | 0.002 | 0.088 | |
| *CCL19* | 0.89 | 9.82 | 0.047 | 0.239 | 0.081 | 0.360 | Chemokines, regulation |
| *CCL3* | 0.89 | 2.84 | 0.036 | 0.229 | 0.105 | 0.360 | Chemokines, regulation |
| *CCL3L1* | 0.91 | 3.48 | 0.036 | 0.229 | 0.105 | 0.360 | Cytokines |
| *CCL4* | 0.91 | 3.62 | 0.036 | 0.229 | 0.105 | 0.360 | Chemokines, regulation |
| *CD3E* | 0.94 | 9.17 | 0.014 | 0.168 | 0.003 | 0.088 | B cell functions, cell functions, T-cell functions |
| *CD5* | 0.97 | 3.93 | 0.014 | 0.168 | 0.003 | 0.088 | B cell functions, |

| | | | | | | | |
|---|---|---|---|---|---|---|---|
| | | | | | | | regulation, T cell functions |
| *CD83* | 0.91 | 2.70 | 0.016 | 0.168 | 0.056 | 0.360 | |
| *CD86* | 1.00 | 2.50 | 0.021 | 0.182 | 0.011 | 0.178 | B cell functions, macrophage functions, regulation, T cell functions |
| *CD97* | 0.86 | 1.47 | 0.004 | 0.130 | 0.002 | 0.088 | |
| *CEACAM1* | 0.86 | 2.26 | 0.041 | 0.234 | 0.146 | 0.413 | Adhesion |
| *CMKLR1* | 0.89 | 3.04 | 0.035 | 0.229 | 0.088 | 0.360 | Chemokines |
| *COL3A1* | 0.89 | 6.36 | 0.022 | 0.182 | 0.038 | 0.322 | Regulation |
| *CXCL1* | 0.91 | 3.25 | 0.041 | 0.234 | 0.146 | 0.413 | Chemokines, regulation |
| *CXCL12* | 0.89 | 2.48 | 0.001 | 0.130 | 0.000 | 0.088 | Chemokines |
| *CYFIP2* | 0.86 | 1.76 | 0.096 | 0.306 | 0.011 | 0.178 | |
| *EGR2* | 0.86 | 1.95 | 0.001 | 0.130 | 0.000 | 0.088 | Regulation |
| *F13A1* | 1.00 | 5.09 | 0.014 | 0.168 | 0.003 | 0.088 | Cell functions |
| *FCGR2A* | 0.94 | 2.30 | 0.014 | 0.168 | 0.003 | 0.088 | Transporter functions |
| *FOS* | 0.94 | 2.27 | 0.016 | 0.168 | 0.105 | 0.360 | |
| *HLA-DRA* | 1.00 | 2.75 | 0.014 | 0.168 | 0.003 | 0.088 | Antigen processing |
| *HLA-G* | 0.97 | 2.22 | 0.004 | 0.130 | 0.002 | 0.088 | Regulation |
| *ICAM2* | 0.97 | 2.01 | 0.021 | 0.182 | 0.011 | 0.178 | Adhesion, regulation |
| *IL10* | 1.00 | 3.64 | 0.010 | 0.168 | 0.006 | 0.104 | Interleukins |
| *IL10RA* | 0.97 | 2.57 | 0.010 | 0.168 | 0.006 | 0.104 | Cytokines |
| *IL12RB2* | 0.86 | 2.01 | 0.114 | 0.350 | 0.016 | 0.241 | Cytokines, NK cell functions, T cell functions |
| *IL18* | 0.91 | 2.41 | 0.005 | 0.130 | 0.023 | 0.270 | Interleukins, NK cell functions, T cell functions |
| *IL1B* | 0.86 | 3.23 | 0.016 | 0.168 | 0.056 | 0.360 | Chemokines, cytokines, interleukins, pathogen defense, regulation |
| *IL34* | 1.00 | 2.85 | 0.014 | 0.168 | 0.003 | 0.088 | Interleukins |
| *IL6* | 0.86 | 5.00 | 0.016 | 0.168 | 0.056 | 0.360 | Interleukins |
| *IL7* | 1.00 | 2.94 | 0.014 | 0.168 | 0.003 | 0.088 | Interleukins |
| *IL7R* | 0.94 | 4.13 | 0.014 | 0.168 | 0.003 | 0.088 | Cytokines |
| *IL8* | 0.89 | 3.14 | 0.036 | 0.229 | 0.105 | 0.360 | Chemokines, cytokines, interleukins, Pathogen defense, |

| | | | | | | | |
|---|---|---|---|---|---|---|---|
| | | | | | | | regulation |
| *IRF2* | 0.89 | 1.33 | 0.004 | 0.130 | 0.002 | 0.088 | Chemokines, regulation |
| *ITGA1* | 0.94 | 1.76 | 0.014 | 0.168 | 0.003 | 0.088 | Adhesion, NK cell functions, T cell functions |
| *ITK* | 0.86 | 4.96 | 0.047 | 0.239 | 0.081 | 0.360 | |
| *LAMP3* | 0.97 | 3.65 | 0.014 | 0.168 | 0.003 | 0.088 | Cell functions |
| *LCK* | 0.89 | 5.92 | 0.047 | 0.239 | 0.081 | 0.360 | Regulation, T cell functions |
| *LILRA1* | 0.89 | 2.21 | 0.035 | 0.229 | 0.088 | 0.360 | Regulation |
| *MAP3K5* | 0.86 | 1.75 | 0.041 | 0.234 | 0.134 | 0.406 | |
| *MAP4K2* | 0.86 | 1.28 | 0.041 | 0.234 | 0.146 | 0.413 | |
| *MEF2C* | 0.89 | 1.64 | 0.041 | 0.234 | 0.016 | 0.241 | |
| *MPPED1* | 0.86 | 2.27 | 0.004 | 0.130 | 0.006 | 0.104 | Cell functions |
| *NEFL* | 0.86 | 3.52 | 0.072 | 0.306 | 0.026 | 0.290 | Cell functions |
| *NLRP3* | 0.86 | 2.28 | 0.005 | 0.130 | 0.023 | 0.270 | |
| *NOTCH1* | 0.97 | 0.65 | 0.004 | 0.130 | 0.002 | 0.088 | Regulation |
| *PDCD1* | 0.97 | 2.51 | 0.014 | 0.168 | 0.003 | 0.088 | Regulation |
| *PECAM1* | 1.00 | 2.18 | 0.014 | 0.168 | 0.003 | 0.088 | Transporter functions |
| *PSMB8* | 0.91 | 1.58 | 0.004 | 0.130 | 0.002 | 0.088 | Chemokines |
| *PTGS2* | 0.91 | 4.64 | 0.036 | 0.229 | 0.105 | 0.360 | Cytokines |
| *PTPRC* | 0.97 | 2.72 | 0.014 | 0.168 | 0.003 | 0.088 | B cell functions, T cell functions |
| *SIGIRR* | 0.89 | 2.44 | 0.016 | 0.168 | 0.053 | 0.360 | |
| *SMPD3* | 0.89 | 1.39 | 0.014 | 0.168 | 0.003 | 0.088 | Cell functions |
| *STAT4* | 0.94 | 3.05 | 0.004 | 0.130 | 0.006 | 0.104 | Chemokines, regulation, T cell functions |
| *TAP1* | 0.86 | 1.58 | 0.004 | 0.130 | 0.002 | 0.088 | Antigen processing |
| *THBD* | 0.89 | 2.17 | 0.010 | 0.168 | 0.006 | 0.104 | Leukocyte functions |
| *TICAM2* | 0.97 | 1.31 | 0.010 | 0.168 | 0.006 | 0.104 | |
| *TLR1* | 0.86 | 1.84 | 0.035 | 0.229 | 0.088 | 0.360 | Microglial functions, TLR |
| *TNFRSF18* | 0.97 | 2.76 | 0.001 | 0.130 | 0.000 | 0.088 | TNF superfamily |
| *TNFRSF1B* | 0.89 | 2.86 | 0.005 | 0.130 | 0.023 | 0.270 | Chemokines, TNF superfamily |
| *TNFSF18* | 0.94 | 2.92 | 0.016 | 0.168 | 0.105 | 0.360 | B cell functions, cell functions, T cell functions, TNF superfamily |
| *TNFSF4* | 0.94 | 1.75 | 0.010 | 0.168 | 0.006 | 0.104 | Chemokines, TNF superfamily |

AUC; Area Under a ROC Curve), Fold change (responder/non-responder), OS; overall survival, PFS: progression-free survival.

**Supplementary Table 5. The prediction performance of random forest models.**

| Random forest model | OOB error rate | Sensitivity | Specificity | # of genes | Gene list |
|---|---|---|---|---|---|
| TNF superfamily | 0.0% | 1.00 | 1.00 | 3 | *IL12RB2, TNFRSF18, TNFSF4* |
| Regulators of T cell activation | 8.3% | 0.80 | 1.00 | 2 | *CD86, LCK* |
| Antigen processing | 8.3% | 1.00 | 0.86 | 2 | *HLA-DRA, TAP1* |
| Adaptive immune response | 8.3% | 1.00 | 0.86 | 11 | *CD3E, CD5, CD86, HLA-DRA, IL12RB2, IL7, IL7R, ITK, PDCD1, STAT4, TAP1* |
| Innate immune response | 8.3% | 0.80 | 1.00 | 14 | *APP, C3, CASP1, CYFIP2, FOS, IL1B, IL34, IL8, MAP3K5, MAP4K2, NLRP3, SIGIRR, TICAM2, TLR1* |
| Inflammation | 16.7% | 0.80 | 0.86 | 4 | *IL1B, IL18, NOTCH1, TLR1* |
| Cytokines | 16.7% | 0.80 | 0.86 | 6 | *CCL3L1, IL10RA, IL12RB2, IL1B, IL7R, IL8* |
| Interleukins | 16.7% | 0.60 | 1.00 | 7 | *IL10, IL18, IL1B, IL34, IL6, IL7, IL8* |
| Adhesion | 25.0% | 0.60 | 0.86 | 3 | *CEACAM1, ICAM2, ITGA1* |
| NK cell functions | 25.0% | 0.60 | 0.86 | 3 | *IL12RB2, IL18, ITGA1* |
| B cell functions | 25.0% | 0.60 | 0.86 | 5 | *CD3E, CD5, CD86, PTPRC, TNFSF18* |
| Humoral immune response | 25.0% | 0.60 | 0.86 | 6 | *CCL3, CD83, IL6, IL7, MEF2C, PDCD1* |
| Inflammasome pathway | 25.0% | 0.60 | 0.86 | 7 | *CASP1, NLRP3, IL18, IL1B, NOTCH1, TLR1, TICAM2* |
| T cell functions | 25.0% | 0.60 | 0.86 | 10 | *CD3E, CD5, CD86, IL12RB2, IL18, ITGA1, LCK, PTPRC, STAT4, TNFSF18* |
| Chemokines | 25.0% | 0.80 | 0.71 | 13 | *CCL19, CCL3, CCL4, CMKLR1, CXCL1, CXCL12, IL1B, IL8, IRF2, PSMB8, STAT4, TNFRSF1B, TNFSF4* |
| All 64 genes | 25.0% | 0.60 | 0.86 | 64 | *APP, C3, CASP1, CCL19, CCL3, CCL3L1, CCL4, CD3E, CD5, CD83, CD86, CD97, CEACAM1, CMKLR1, COL3A1, CXCL1, CXCL12, CYFIP2, EGR2, F13A1, FCGR2A, FOS, HLA-DRA, HLA-G, ICAM2, IL10, IL10RA, IL12RB2, IL18, IL1B, IL34, IL6, IL7, IL7R,* |

| | | | | | |
|---|---|---|---|---|---|
| | | | | | *IL8, IRF2, ITGA1, ITK, LAMP3, LCK, LILRA1, MAP3K5, MAP4K2, MEF2C, MPPED1, NEFL, NLRP3, NOTCH1, PDCD1, PECAM1, PSMB8, PTGS2, PTPRC, SIGIRR, SMPD3, STAT4, TAP1, THBD, TICAM2, TLR1, TNFRSF18, TNFRSF1B, TNFSF18, TNFSF4* |

OOB error rate; Out-of-bag error rate.

**Supplementary Table 6. The mean number of immune cells per mm$^2$ in tumor by immunohistochemistry.**

| Patients | Response | CD3 (cells/mm$^2$) | CD8 (cells/mm$^2$) | CD68 (cells/mm$^2$) |
|---|---|---|---|---|
| A1 | R | 12.4 | 42.47 | 495.68 |
| A2 | NR | 26.5 | 10.09 | 585.23 |
| A3 | NR | 43.9 | 40.07 | 443.79 |
| A4 | NR | Not available | 12.28 | 56.77 |
| A5 | NR | 15.8 | 3.15 | 138.8 |
| A6 | R | 786.3 | 717.65 | 1762.98 |
| A7 | R | 72.3 | 54.07 | 657.44 |
| A8 | NR | 28.5 | 6.88 | 164.01 |
| A10 | NR | 28.6 | 18.82 | 410.9 |
| A11 | R | 13.9 | 19.63 | 106.15 |
| A12 | NR | 32.3 | 11.02 | 156.8 |
| A14 | R | 29.0 | 14.04 | 83.6 |

R; Responder, NR; Non-responder.
